# Increased Risk of Cirrhosis in Patients with Inflammatory Bowel Disease: A Danish registry-based cohort study (1998-2018)

**DOI:** 10.1101/2024.03.05.24303668

**Authors:** Parakkal Deepak, Scott McHenry, Anastasia Karachalia Sandri, Maiara Brusco De Freitas, Mohammad Zamani, Andres J. Yarur, Tine Jess

## Abstract

**Background and Aims:** Prior studies suggest an increased risk of non-alcoholic fatty liver disease (NAFLD) in patients with inflammatory bowel disease (IBD). We aimed to investigate the risk of cirrhosis in a nationwide cohort of IBD patients compared to a matched non-IBD population.

**Methods:** Patients diagnosed with IBD without prior cirrhosis during 1998-2018 were identified in the Danish health registries and were matched 1:10 to persons without IBD. Cox regression was used to calculate hazard ratios (HRs) with corresponding 95% confidence intervals (CIs).

**Results:** Within the study population of 495,220 persons, a total of 2,741 cirrhosis cases were identified during follow-up, with a higher proportion of cases among patients with IBD (0.9%) compared to non-IBD persons (0.5%). Patients with IBD had a significantly higher risk of cirrhosis compared to non-IBD persons (adjusted HR (aHR) (95% CI): 1.84 (1.64-2.04)). The leading etiology of cirrhosis in IBD was NAFLD (51.6%), followed by alcohol (39.0%). The risk of cirrhosis among IBD patients (compared to non-IBD persons) was more pronounced among those diagnosed with IBD ≤ 40 years of age (aHR (95% CI): 3.08 (2.45-3.87); vs. > 40 years of age, 1.63 (1.45-1.84); p-value <0.001) and CD patients (aHR (95% CI): 2.20 (1.80-2.67); vs. 1.72 (1.52-1.95) among UC; p-value 0.04).

**Conclusion:** IBD was associated with an increased risk of incident cirrhosis, especially in patients aged ≤ 40 years at IBD diagnosis and in patients with CD. These findings point towards a need for focused screening for cirrhosis among IBD patients, especially in certain groups.

## Introduction

Obesity and cardiometabolic disorders are increasingly observed comorbidities in patients with inflammatory bowel disease (IBD) despite the common characterization of IBD as a disease of diarrhea, malnutrition, and wasting^1^. Of particular importance within the cardiometabolic spectrum of disorders in IBD is the association with non-alcoholic fatty liver disease (NAFLD). Up to one-third of patients with IBD develop NAFLD worldwide, with a risk of NAFLD two times higher compared to healthy subjects^2^. The link between IBD and NAFLD is important since it predisposes to both cardiometabolic and liver-related complications such as hypertension, dyslipidemia, diabetes mellitus, cirrhosis, and hepatocellular carcinoma^3^.

While the association between NAFLD and IBD has been characterized across many studies, the progression to advanced liver fibrosis and cirrhosis is less clear since prior reports exploring the progression have been limited to retrospective, cross-sectional, and single-center cohort studies^4–6^. An increase in mortality related to NAFLD in patients with IBD has also been reported in a meta-analysis of all-cause and cause-specific mortality, where it is unclear if this is driven by a progression to liver cirrhosis and resultant complications and if this observation can be reproduced when studied in a longitudinal population-based and hence unselected study design^7^.

Thus, there is a critical need to better understand the risk of progression to liver cirrhosis in patients with IBD and if this is driven by the previously noted association with NAFLD. Our aim was to investigate the risk of cirrhosis (compensated or decompensated) in persons with IBD when compared to a matched non-IBD population using the Danish nationwide health registries.

## Methods

### Study design

This is a nationwide registry-based cohort study from Denmark using prospectively collected data. Eligible individuals with a diagnosis of IBD were matched 1:10 to non-IBD persons.

### Data sources

The data sources included the National Patient Register (NPR), the National Register of Pathology (NRP), the Danish Civil Registration System (CPR), and the Danish Cause of Death Register (DAR) as detailed in *supplementary methods*.

### Study period

The study period was from January 1, 1998 until December 31, 2018.

### Study cohort

#### IBD population

Newly diagnosed persons with IBD (incident cases) were identified between (and including) 1998-2018 using the International Classification of Diseases (ICD) codes in the NPR as detailed in *supplementary methods*. For eligible IBD persons, the date of the first IBD registration was considered to be the ‘exact’ date of IBD diagnosis.

#### Matched non-IBD population

Each person in the IBD population was matched to ten non-IBD individuals using the second IBD registration as the matching date as detailed in *supplementary methods*. Matching was performed on the basis of sex (at birth), age (in six-month intervals), year of IBD diagnosis (in six-month intervals), and the municipality of residence (‘*kommune’* in Danish) at the matching date.

### Study outcome

The study outcome was a diagnosis of liver cirrhosis, including both compensated and decompensated cirrhosis, as recorded by relevant ICD-8/10 or SNOMED codes (**Appendix Table A1**)^8, 9^.

#### Potential etiology and method of diagnosis of liver cirrhosis

For persons in the study cohort who received a diagnosis of cirrhosis during the follow-up period, we further examined for etiology as detailed in *supplementary methods* and **Appendix Table A2** and examined the method of diagnosis of liver cirrhosis when available, that is through biopsy and/or imaging (**Appendix Table A3**).

### Statistical analyses

Time-to-event analysis was performed with individuals followed from the IBD diagnosis/index date (for the IBD and non-IBD population, respectively) until the diagnosis of liver cirrhosis (study outcome), death, emigration, or end of follow-up (December 31, 2018), whichever came first. Persons in the matched non-IBD population were additionally censored in case of an IBD registration during follow-up. The Kaplan-Meier estimator was utilized to calculate the overall cumulative incidence of cirrhosis among persons with IBD, compared to the non-IBD population but also stratified by type of IBD (UC/CD), sex (female/male), and age group at IBD diagnosis/index date (≤40; >40 years of age). Cox proportional regression analysis was performed to calculate hazard ratios (HRs) with corresponding 95% confidence intervals (CIs) for cirrhosis among persons with IBD, compared to the non-IBD population. Adjusted analysis was also performed for the type of IBD, sex, age group, and year of study entry.

### Sensitivity analysis

Time-to-event analysis was performed for the period 2002-2018, where we extended the primary study outcome (diagnosis of liver cirrhosis) to include liver-related death too (composite study outcome) (*supplementary methods* and **Appendix Table A4**).

## Results

A total of 63,480 newly diagnosed persons with IBD (incident cases) were identified in the Danish registries, out of whom 61,524 were classified as UC or CD, with 46,385 diagnosed during the study period (1998-2018) (**Figure 1**). Of these, 45,020 patients met the residency criteria and had no diagnosis of liver cirrhosis prior to their IBD diagnosis. These individuals were matched 1:10 to a non-IBD population resulting in a total study population of 495,225 persons (**Figure 1**).

**Figure 1.**
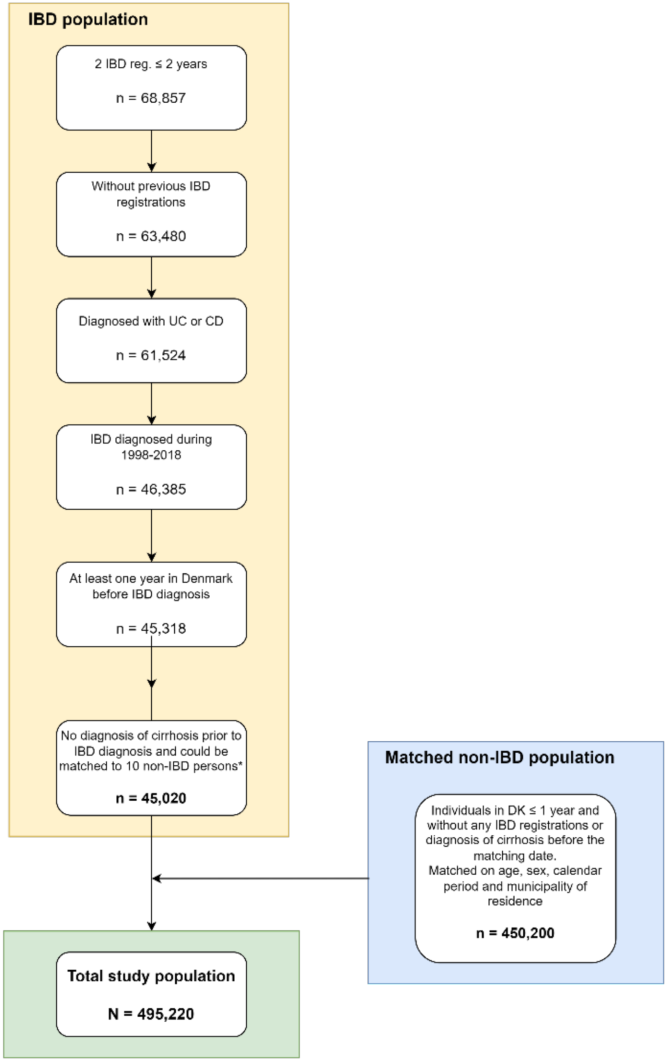
Flowchart of study population *n=288 had a diagnosis of cirrhosis; n=10 could not be matched to 10 non-IBD persons (had fewer than 10 matches each).

The characteristics (IBD type, sex, age group at IBD diagnosis/index date and year of cohort entry) of the entire study population along with the number of cirrhosis cases are shown in **Table 1**. This information is also presented for the IBD cohort and the matched non-IBD individuals separately (**Table 1**). In the study population of 495,225 individuals, a total of 2,741 (0.55%) persons received a diagnosis of liver cirrhosis during follow-up, out of whom 410 (0.91%) were in the IBD group and 2,331 (0.52%) were in the non-IBD group. The median age at the time of cirrhosis was 54 years (inter-quartile range (IQR), 41-64) in the IBD group and 56 (IQR, 46-66) in the non-IBD group. Among those in the study population who got a diagnosis of cirrhosis during follow up, the median time (in years) from IBD diagnosis/index date to diagnosis of cirrhosis was 5.0 years (IQR, 1.2-9.8) among persons with IBD, and 6.4 (IQR, 3.2-10.8) within the non IBD population.

**Table 1.** Characteristics of the study population and number of cirrhosis cases.

|  |  | **Total no. of persons (C %)** | **Total no. of persons with cirrhosis (R %)** | **No. of persons with IBD** | **No. of persons with cirrhosis among persons with IBD (R %)** | **No. of persons without IBD** | **No. of persons with cirrhosis among persons without IBD (R %)** |
| --- | --- | --- | --- | --- | --- | --- | --- |
|  |  | N=495,220 | N=2,741 | N=45,020 | N=410 | N=450,200 | N=2,331 |
| IBD type | UC | 340,670 (68.6) | 2,033 (0.6) | 30,970 | 290 (0.9) | 309,700 | 1,743 (0.6) |
|  | CD | 154,550 (31.2) | 708 (0.5) | 14,050 | 120 (0.8) | 140,500 | 588 (0.4) |
| Sex | Female | 264,264 (53.4) | 1,208 (0.5) | 24,024 | 187 (0.8) | 240,240 | 1,021 (0.4) |
|  | Male | 230,956 (46.6) | 1,533 (0.7) | 20,996 | 223 (1.1) | 209,960 | 1,310 (0.6) |
| Age group (in years) at IBD diagnosis/index date | ≤ 16 | 29,238 (5.9) | 20 (0.1) | 2,658 | 13 (0.5) | 26,580 | 7 (<0.1) |
|  | [17-40] | 159,709 (32.3) | 394 (0.2) | 19,959 | 84 (0.4) | 199,590 | 310 (0.2) |
|  | [41-64] | 219,549 (44.3) | 1,589 (1.0) | 14,519 | 220 (1.5) | 145,190 | 1,369 (0.9) |
|  | ≥ 65 | 86,724 (17.5) | 738 (0.8) | 7,884 | 93 (1.2) | 78,840 | 645 (0.8) |
| Mean age (SD) |  | 42.8 (19.5) | 54.9 (14.3) | 42.8 (19.5) | 51.4 (16.3) | 42.8 (19.5) | 55.6 (13.8) |
| Median age (IQR) |  | 40 (26-58) | 56 (45-65) | 40 (26-58) | 54 (41-64) | 40 (26-58) | 56 (46-66) |
| Year of cohort entry (IBD diagnosis year/index year) | 1998-2001 | 74,745 (15.1) | 794 (1.1) | 6,795 | 90 (1.3) | 67,950 | 704 (1.0) |
|  | 2002-2005 | 89,298 (18.0) | 776 (0.9) | 8,118 | 123 (1.5) | 81,180 | 653 (0.8) |
|  | 2006-2009 | 94,116 (19.0) | 612 (0.7) | 8,556 | 95 (1.1) | 85,560 | 517 (0.6) |
|  | 2010-2013 | 102,014 (20.6) | 378 (0.4) | 9,274 | 62 (0.7) | 92,740 | 316 (0.3) |
|  | 2014-2018 | 135,047 (27.3) | 181 (0.1) | 12,277 | 40 (0.3) | 122,770 | 141 (0.1) |

IBD: inflammatory bowel disease; IQR: interquartile range; C: column; R: row; SD: standard deviation.

### Risk of cirrhosis in IBD persons compared to non-IBD population

The cumulative incidence of cirrhosis during follow-up was significantly higher within the IBD cohort compared to the matched non-IBD group (p-value < .0001) (**Figure 2a**). After adjusting for the type of IBD, sex, age group, and year of study entry, IBD persons were at a significantly higher hazard for liver cirrhosis compared to non-IBD individuals (aHR, 1.84; 95% CI, 1.65 - 2.04) (**Figure 3**). A significant difference was also noted when exploring UC and CD separately (p-value < .0001) (**Figure 2b**). In the Cox proportional regression model, patients with CD had a significantly higher risk for a cirrhosis compared to their matched non-IBD population (aHR, 2.20; 95% CI, 1.80 - 2.67), whereas the risk was slightly lower, although still increased, in UC (aHR, 1.72; 95% CI, 1.52 - 1.95, p-value = 0.04) (**Figure 3**). On the Kaplan-Meier analysis, a significant difference was seen among the survival curves for cirrhosis comparing males and females to their corresponding matched non-IBD population (p-value < .0001) (**Figure 2c**); however, no significant differences were noted between males and females in the adjusted Cox model (p-value = 0.42) (**Figure 3**). Regarding age group at IBD diagnosis/index date, a significant difference was again among the survival curves for cirrhosis comparing those ≤ 40 years of age and >40 to their corresponding matched non-IBD population (p-value < .0001) (**Figure 2d**). Persons up to 40 years of age at IBD diagnosis had a threefold higher risk of cirrhosis compared to non-IBD individuals (aHR, 3.08; 95% CI, 2.45-3.87), while that risk was lower yet still significant among persons older than 40 years of age at the time of IBD diagnosis (aHR, 1.63; 95% CI, 1.45-1.84) (**Figure 3**). Additionally, an increasing HR for liver cirrhosis in IBD patients was demonstrated over calendar time, increasing from 1.37 (95% CI: 1.07-1.66) during 1998-2001 to 2.89 (95% CI: 2.03-4.10) during 2014-2018 (**Figure 3**).

**Figure 2.**
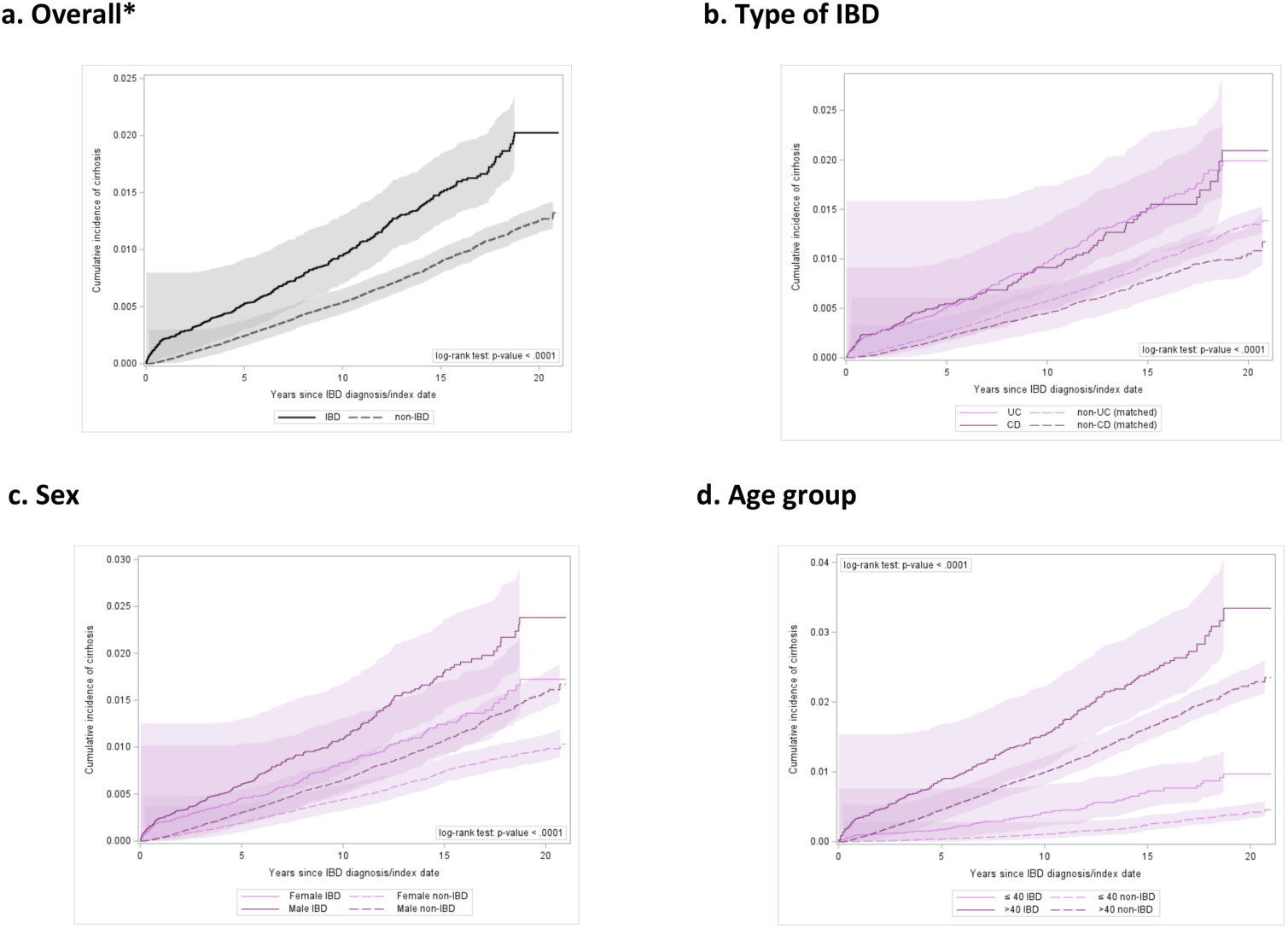
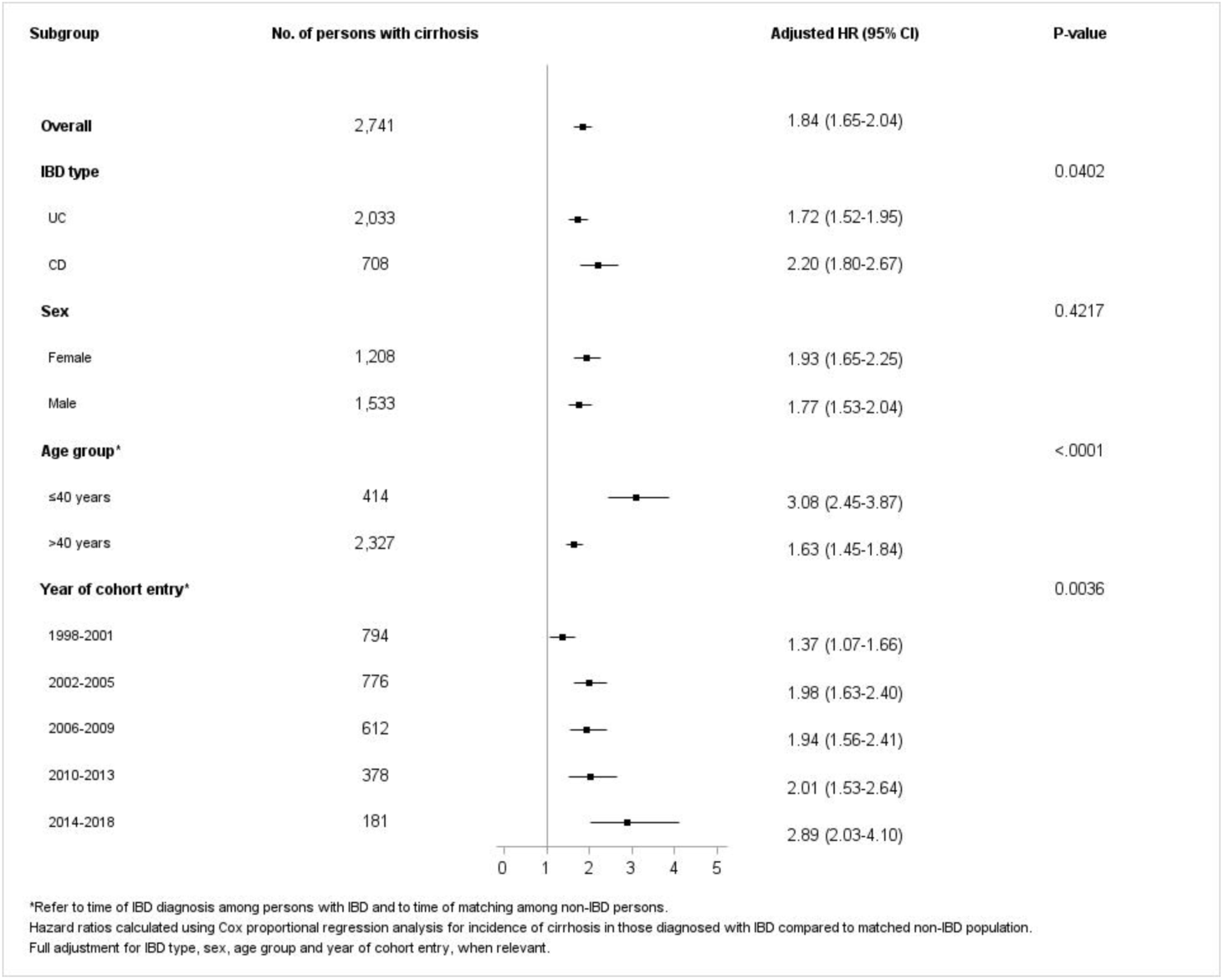
Kaplan-Meier survival analysis curves for cumulative incidence of cirrhosis *Event/Censoring outcomes (i) in the entire study population: 2,741 cirrhosis; 44,530 death; 10,725 emigration; 437,224 end of follow-up without cirrhosis; (ii) in IBD population: 410 cirrhosis; 4,784 death; 1,070 emigration; 38,756 end of follow-up without cirrhosis; (iii) in non-IBD population: 2,331 cirrhosis; 39,746 death; 9,665 emigration; 398,468 end of follow-up without cirrhosis.

**Figure 3.**
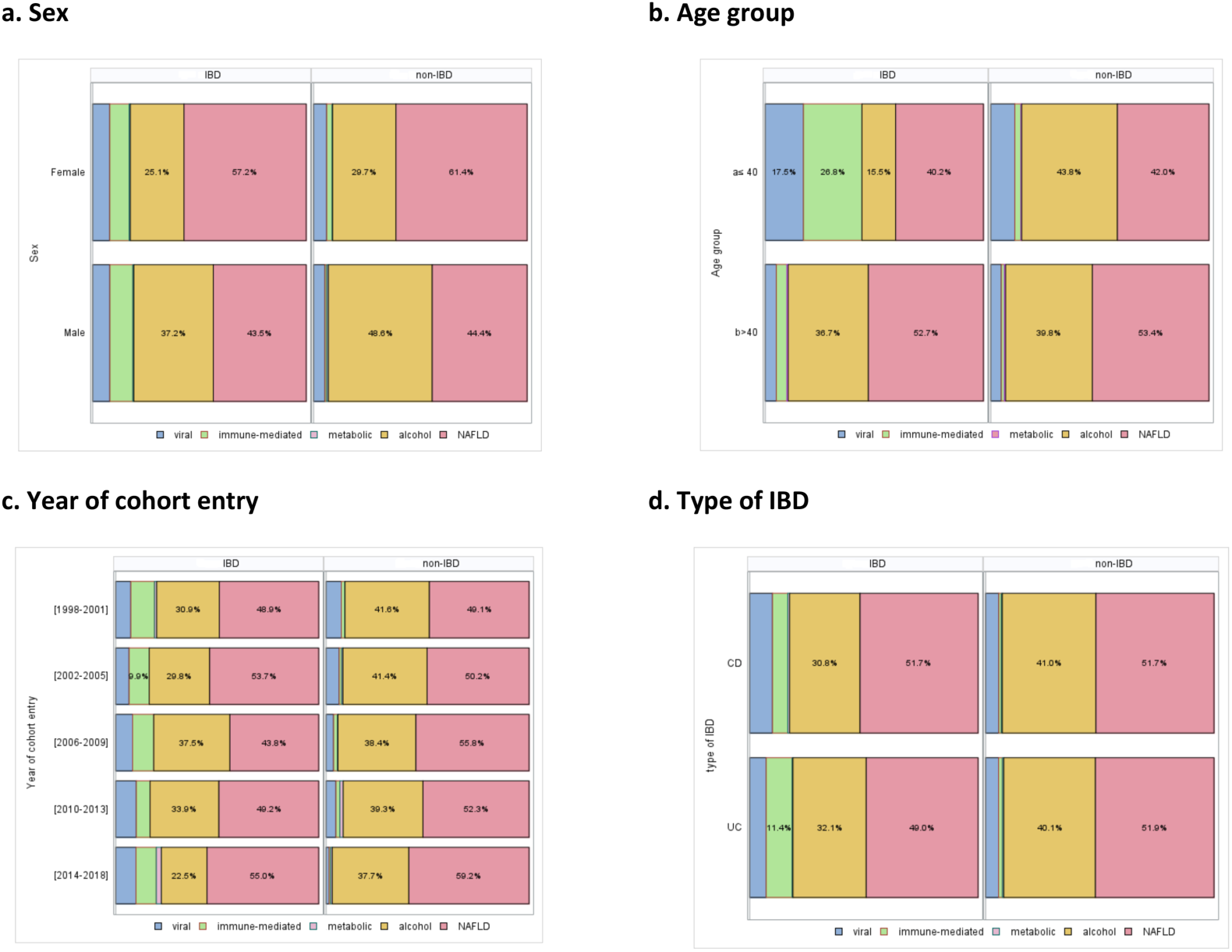
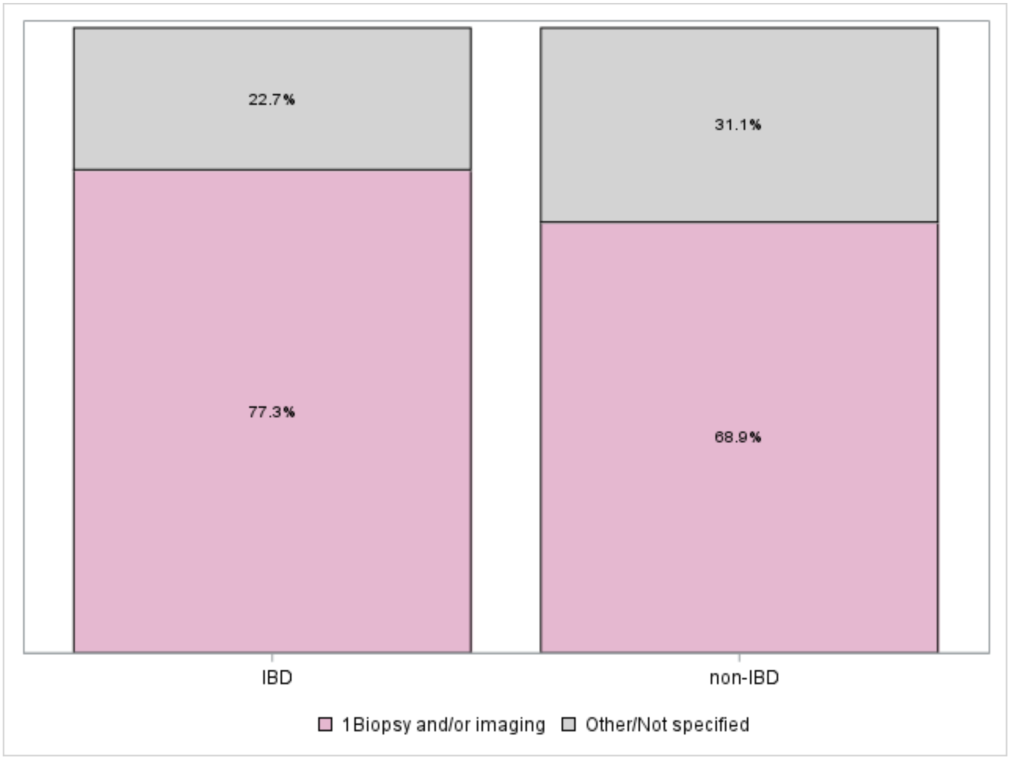
Forest plot of adjusted HR (95% CI) for cirrhosis in persons with IBD compared to matched non-IBD population in Denmark 1998-2018

### Etiology of liver cirrhosis and method of diagnosis

For persons in the study population who received a diagnosis of cirrhosis during the follow-up period (N = 2,741), we further examined for any previous registration(s) of viral hepatitis, immune-mediated, primary metabolic, alcohol-related, or cryptogenic/NAFLD liver disease, as the underlying etiology of cirrhosis. The most common primary registration was NAFLD identified in 51.6% of persons, followed by alcohol-related registration in 39.0% of persons (**Appendix Table A5**). We found that females had a greater percentage of NAFLD compared to males, both in the IBD group (57.2% vs. 43.5%) and the non-IBD group (61.4% vs. 44.4%) (**Figure 4**). The year of cohort analysis for etiology showed that the percentage of persons with NAFLD as leading registration was similar in the IBD and the matched non-IBD populations over the years, reaching a high of 55% in the IBD persons and 59.2% in the non-IBD populations in 2014-2018. NAFLD as the leading etiology for liver cirrhosis was comparable in the study population for IBD type (51.7% in CD and 49.0% in UC) and in their matched non-IBD populations (**Figure 4**).

**Figure 4.**
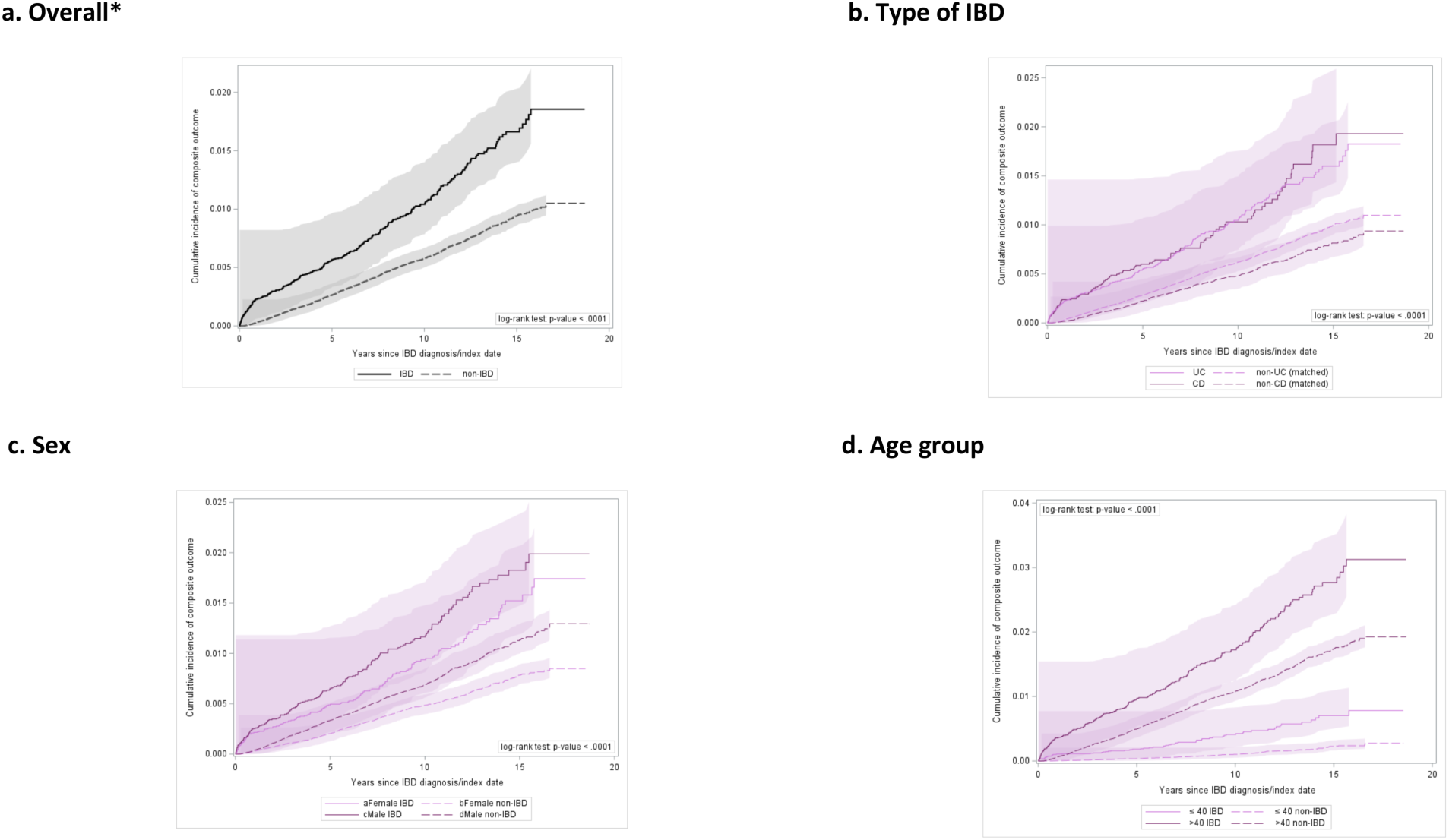
Etiology of cirrhosis in the study population (N=2,741).

Liver cirrhosis was diagnosed based on biopsy and/or imaging for a total of 1,924 individuals with cirrhosis (70.2%) in the study population. Comparable proportions were noted in terms of biopsy and/or imaging for the IBD and the non-IBD group, with the former having a somewhat higher percentage than the latter (77.3% vs. 68.9%, respectively (**Supplementary Figure 1**).

### Sensitivity Analysis

Within the study population of 420,475 persons (38,225 IBD persons, along with their 382,250 controls), a total of 2,064 events (composite study outcome) took place during follow-up, that is 1,947 diagnoses of liver cirrhosis and 117 liver-related deaths. Out of these, 329 were among persons with IBD (320 cases of cirrhosis and 9 liver-related deaths), and the remaining 1,735 were among persons without IBD (1,627 cases of cirrhosis and 108 liver-related deaths). Sensitivity analyses results using the Kaplan-Meier estimator were very similar to those of the primary analyses. The cumulative incidence of liver cirrhosis/liver-related death during follow-up was significantly higher in the IBD group compared to the matched non-IBD population not only as a total but also when stratified by type of IBD, sex, and age groups (all p-values < .0001) **(Supplementary Figure 2**). The results from Cox regression were consistent with those of the primary analyses, even though the estimates now were slightly higher. Specifically, after adjusting for the type of IBD, sex, age group, and year of study entry, persons with IBD remained at a significantly higher hazard for developing cirrhosis/liver-related death compared to the matched non-IBD group (aHR, 1.98; 95% CI, 1.76 - 2.23) (**Table 2**).

**Table 2.**
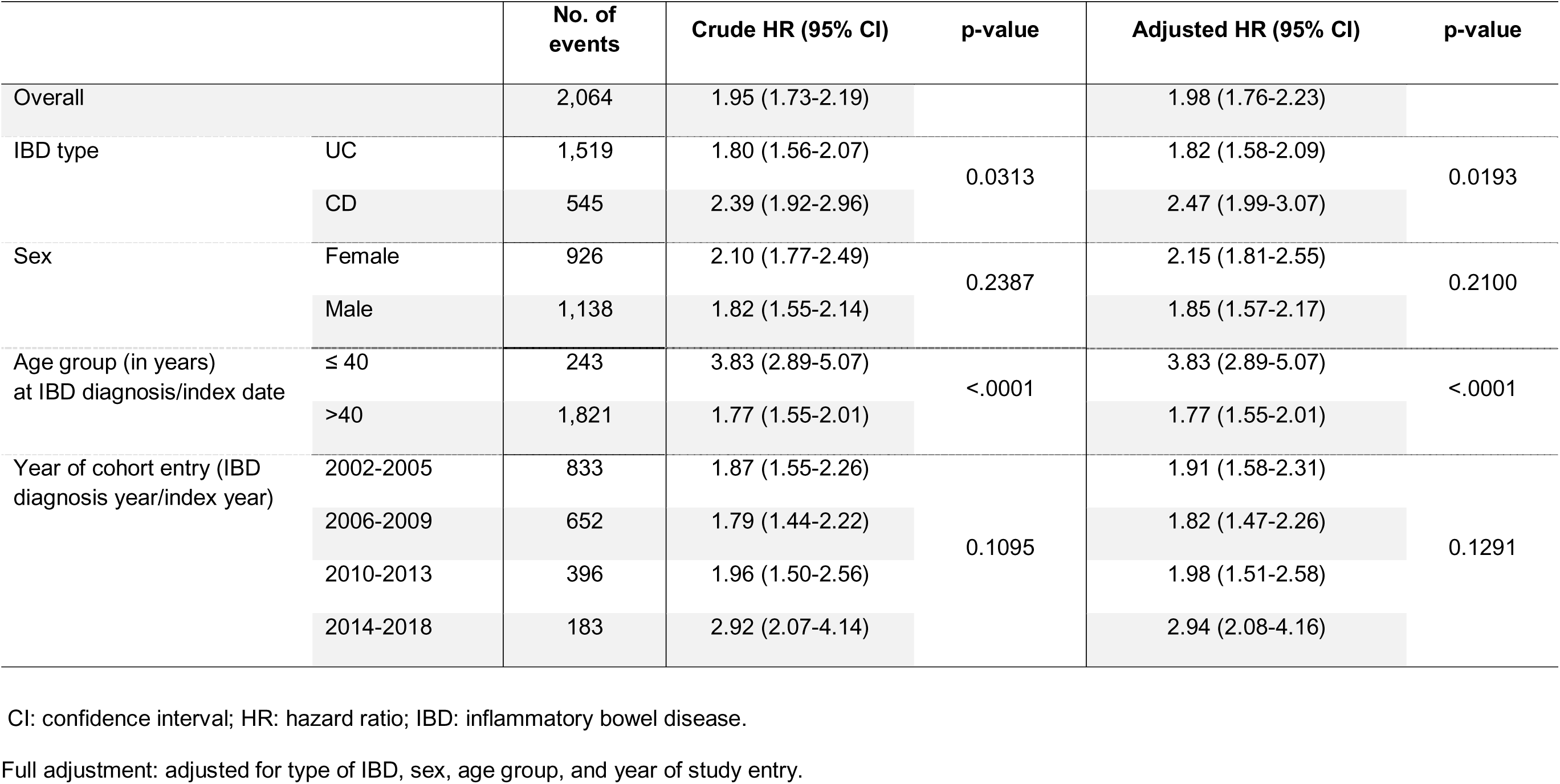
Hazard ratio for cirrhosis or liver-related death (composite outcome) in IBD compared to matched non-IBD population in Denmark 2002-2018.

|  |  | No. of events | Crude HR (95% CI) | p-value | Adjusted HR (95% CI) | p-value |
| --- | --- | --- | --- | --- | --- | --- |
| Overall |  | 2,064 | 1.95 (1.73-2.19) |  | 1.98 (1.76-2.23) |  |
| IBD type | UC | 1,519 | 1.80 (1.56-2.07) | 0.0313 | 1.82 (1.58-2.09) | 0.0193 |
|  | CD | 545 | 2.39 (1.92-2.96) |  | 2.47 (1.99-3.07) |  |
| Sex | Female | 926 | 2.10 (1.77-2.49) | 0.2387 | 2.15 (1.81-2.55) | 0.2100 |
|  | Male | 1,138 | 1.82 (1.55-2.14) |  | 1.85 (1.57-2.17) |  |
| Age group (in years)<br>at IBD diagnosis/index date | ≤ 40 | 243 | 3.83 (2.89-5.07) | <.0001 | 3.83 (2.89-5.07) | <.0001 |
|  | >40 | 1,821 | 1.77 (1.55-2.01) |  | 1.77 (1.55-2.01) |  |
| Year of cohort entry (IBD<br>diagnosis year/index year) | 2002-2005 | 833 | 1.87 (1.55-2.26) | 0.1095 | 1.91 (1.58-2.31) | 0.1291 |
|  | 2006-2009 | 652 | 1.79 (1.44-2.22) |  | 1.82 (1.47-2.26) |  |
|  | 2010-2013 | 396 | 1.96 (1.50-2.56) |  | 1.98 (1.51-2.58) |  |
|  | 2014-2018 | 183 | 2.92 (2.07-4.14) |  | 2.94 (2.08-4.16) |  |

CI: confidence interval; HR: hazard ratio; IBD: inflammatory bowel disease.
Full adjustment: adjusted for type of IBD, sex, age group, and year of study entry.

## Discussion

The association between NAFLD and IBD is now well recognized^2, 10^; however, if this association translates into an increased risk of developing cirrhosis is not currently known. This distinction is important as only a minority of NAFLD patients will have nonalcoholic steatohepatitis (NASH)^11^ while those with simple steatosis are not considered at a particularly high risk of developing liver-related complications^12^. In this population-based cohort study including 45,020 persons with IBD and 450,200 matched non-IBD persons, we have shown a nearly 2-fold increased risk of incident liver cirrhosis and liver-related death in the IBD population.

Almost all (~90%) of the IBD patients that developed cirrhosis fit under the new umbrella of steatotic liver disease (SLD)^13^ with nearly half being NAFLD (51.6%) and almost all the remaining being alcohol-related (39.0%). This increased risk of cirrhosis among IBD patients was more pronounced when they were diagnosed with IBD before 40 years of age and in those with Crohn’s disease. This increased risk is becoming even more pronounced in the modern era despite improved IBD care pathways.

Our study findings, showing an elevated and accelerated risk of cirrhosis and liver-related death in persons with IBD, strengthen the case to screen all IBD patients for hepatic steatosis and cirrhosis, especially in those with IBD diagnosed ≤ 40 years of age and with the IBD subtype of CD. The goal would be to identify early-stage liver disease, potentially preventing its progression to cirrhosis. Those found to have underling liver disease would then be connected both to liver-related and cardiometabolic care. Importantly, prior studies have noted an increased risk of premature atherosclerotic cardiovascular disease in patients with IBD, and that cirrhosis in the non-IBD population has also been noted to increase the risk of adverse cardiovascular outcomes^14^. Considering these facts, preventing cirrhosis becomes critical to potentially reduce the already increased risk of cardio-vascular events in IBD patients. We additionally confirmed the prior observation of increased liver-related mortality in a metaanalysis^7^ in this longitudinal population-based design with a nearly 2-fold increased risk of incident liver cirrhosis and liver-related death.

The underlying pathogenesis explaining the increased risk of NAFLD in patients with IBD remains unclear. We have shown in a prior study that NAFLD in CD is associated with quiescent disease, history of an ileocecal resection, and elevated IL-8 levels in the blood^15^. The leaky-gut hypothesis is often proposed as a mechanism of NAFLD pathogenesis in the general population^16^. Based on this theory, hepatic steatosis is hypothesized to develop secondary to chronic intestinal inflammation, dysbiosis, increased intestinal permeability, translocation of luminal microbial components, and secretion of pro-inflammatory cytokines^16, 17^. Intestinal barrier dysfunction has been shown to be associated with the later development of CD as well as one of the earliest preclinical events in the pathogenesis of CD^18^. This may explain both the higher incidence and accelerated risk of liver cirrhosis as well as the greater risk of cirrhosis in those with IBD diagnosed ≤ 40 years of age, with a longer duration of chronic intestinal inflammation, since barrier dysfunction remains evident in some patients with quiescent Crohn’s disease^19^.

In UC, the mechanism explaining the increased risk of cirrhosis is less clear, although altered tight junction structures contributing to the impaired epithelial barrier function have also been shown in patients with UC^20^. The reasons for a higher risk of cirrhosis in persons with CD compared to UC is also unclear. CD patients with NAFLD are more likely to have a histologic form that is characterized by hepatocellular necroinflammation and NASH^21^. Other postulated mechanisms for increased rates of NAFLD seen include the decreased expression of the ileal farnesoid X receptor (FXR) gene and lower levels of fibroblast growth factor 19 (FGF 19)^22^. Since FXR is predominantly found in the small intestine, this may partly explain why our study found an increased risk of cirrhosis in patients with CD as compared to those with UC. Furthermore, persons with CD are more likely to undergo ileal resections with disruption of bile acid homeostasis or deficiency in FGF 19, whose deficiency has been associated with NAFLD^23^. Moreover, CD is associated with a higher degree of gut microbiome alteration when compared to UC and dysbiosis can be pivotal in the pathway to NAFLD development and progression to cirrhosis^24^.

This study has several strengths. To our knowledge, this is the first study exploring the risk of cirrhosis among persons with IBD compared to non-IBD individuals at a population level. The definition of IBD diagnosis used in this study (that is two IBD registrations up to two years apart) is a validated method for the NPR that renders misclassification of IBD very unlikely^25^. The population-based study design allowed for the inclusion of a sizeable and unselected cohort of around half a million individuals over a 21-year time period, since the Danish CPR system offers complete coverage of the entire population living in Denmark since 1968. We carefully selected 1998 as the start of our study period to ensure complete coverage of liver biopsies (complete coverage in NRP since 1990) and of psychiatric disorders including alcohol-related ones (complete coverage in NPR since 1995) during the follow-up period (i.e.1998-2018). This decision also allowed for complete coverage for some time before study entry (i.e. 1995-1997) to ensure that only eligible individuals were included in the study population. The consistency between the primary and the sensitivity analyses, strengthens the robustness of the study findings, including the at-risk groups and over the years of the study, which potentially speaks to their generalizability. We not only considered the etiology of liver cirrhosis, but also how the diagnosis was established, showing similar trends in the IBD and non-IBD populations. This thereby, minimizes the likelihood that the study findings could be solely explained by surveillance bias.

Despite our best efforts in designing and executing the current study, we do need to consider certain limitations. The etiology of cirrhosis was derived by using a hierarchical strategy according to which NAFLD was identified through a process of ruling out other etiologies and prior to the wide-spread adoption of the SLD paradigm^13^, hence there is the risk of misclassification. However, this approach is similar to that used in the hepatology clinic using a variety of sources and is unlikely to impact the validity of our conclusions considering such misclassification would apply to both the IBD and the non IBD population. Data on body mass index (BMI) is not recorded routinely in the Danish registers so we could not account for it in the analyses. Even though differences in the obesity rates between those with and without IBD, might have been a confounding factor, prior studies in Denmark exploring the association between BMI and IBD have failed to show a linear association with increasing BMI and the obesity prevalence in Denmark has increased over time across all ages and birth cohorts^26, 27^. Hence, it is unlikely that persons with IBD had a significantly higher BMI when compared to the matched non-IBD population. Finally, immortal time bias related to our case definition of IBD requiring two registrations should also be addressed. Even though the two IBD registrations could be up to two years apart, in practice, among the 45,020 eligible IBD persons included in this study, the median time between these two registrations was only 27 days (IQR: 13-57 days). Furthermore, the second IBD registration date was used for matching to ensure balance between the two comparator groups.

In conclusion, we found that patients with IBD have an increased and accelerated risk of liver cirrhosis and liver-related death over a 20-year study period in Denmark among persons with IBD when compared to non-IBD individuals, particularly significant for those diagnosed with IBD at age ≤ 40 years and for persons with Crohn’s disease. This data advocates for the need for screening all patients with IBD for NAFLD and cirrhosis, with higher-risk groups identified within the overall pool of persons with IBD. Future studies looking into the underlying mechanisms of this association and potential strategies to prevent and treat them are warranted.

## Data Availability Statement

Data is from the Danish Health Registry and accessed through the PREDICT center and hence is not available to the other researchers except through direct contact with PREDICT center.

## Authors’ Contributions

PD: Study concept, study design, interpretation of the data, drafting of the initial draft of the manuscript, acquiring funding, and equal supervision of the study.

SM: Study concept, study design, interpretation of the data, and drafting of the initial draft of the manuscript.

AKS: Study design, data extraction, analysis of the data, interpretation of the data, and critical review of the manuscript.

MZ, AY, MBDF: Interpretation of the data, and critical review of the manuscript.

TJ: Study design, interpretation of the data, critical review of the manuscript, and equal supervision of the study.

