## Supplementary Tables for "Increased Risk of Cirrhosis in Patients with Inflammatory Bowel Disease: A Danish registry-based cohort study (1998-2018)"

**Appendix**

**Methods:**

**Data sources**

The data sources included the National Patient Register (NPR), the National Register of Pathology (NRP), the Danish Civil Registration System (CPR), and the Danish Cause of Death Register (DAR). All persons living in Denmark are assigned a unique personal identification number at the time of birth or immigration that enables linkage of information across the registers. The NPR captures all inpatient encounters from January 1, 1977 and all outpatient encounters dating from January 1, 1995. The NRP holds information on all liver biopsies performed in hospital departments in Denmark since January 1, 1990 when electronic SNOMED codes were introduced.

**Study cohort**

*IBD population*

Newly diagnosed persons with IBD (incident cases) were identified between (and including) 1998-2018 using the International Classification of Diseases (ICD) codes (ICD-10 code DK51, and ICD-8 codes 563.19 and 569.04 for ulcerative colitis (UC); ICD-10 code DK50, and ICD-8 codes 563.08 and 563.09 for Crohn’s disease (CD)) in the NPR. An IBD diagnosis was based on at least two IBD registrations up to two years apart, where registration refers to an outpatient visit or to an inpatient contact associated with a primary or secondary IBD registration. For IBD diagnosis to be classified as UC or CD, both IBD registrations had to indicate UC or CD, respectively; persons with discordant registrations (i.e., one as UC and one as CD) were considered to have IBD unclassified (IBD-U) and were excluded from this study. Individuals also had to have been in Denmark for at least one year prior to their first IBD registration (as well as between the two IBD registrations). Persons with a diagnosis of liver cirrhosis prior to their IBD registrations were excluded from the IBD study population. For eligible IBD persons, the date of the first IBD registration was considered to be the ‘exact’ date of IBD diagnosis.

*Matched non-IBD population*

Each person in the IBD population was matched to ten non-IBD individuals using the second IBD registration as the matching date. Matching was performed on the basis of sex (at birth), age (in six-month intervals), year of IBD diagnosis (in six-month intervals), and the municipality of residence (‘*kommune’* in Danish) at the matching date. Candidate non-IBD individuals included persons without any IBD registrations and a diagnosis of liver cirrhosis prior to the matching date. They also had to have been living in Denmark at least one year prior to the matching date (in accordance with the IBD population). Persons of [80-90) years of age were matched as one group, and the same was done for persons ≥ 90 years old.

**Study outcome**

The study outcome was a diagnosis of liver cirrhosis, including both compensated and decompensated cirrhosis, as recorded by relevant ICD-8/10 or SNOMED codes (**Appendix Table A1**)^13, 14^.

*Potential etiology and method of diagnosis of liver cirrhosis*

For persons in the study cohort who received a diagnosis of cirrhosis during the follow-up period, we further examined for any previous registration(s) of viral hepatitis, immune-mediated, primary metabolic, alcohol-related, or cryptogenic/NAFLD liver disease. Relevant ICD-8/10 and SNOMED codes were once again used in this case (**Appendix Table A2**). For persons who had registrations in more than one of those disease categories (such as immune-mediated and alcohol-related), we used a hierarchy of: viral hepatitis > immune-mediated > primary metabolic > alcohol-related > cryptogenic/NAFLD liver disease. We also examined the method of diagnosis of liver cirrhosis when available, that is through biopsy and/or imaging (**Appendix Table A3**), by considering a window of

**Sensitivity analysis**

Time-to-event analysis was performed where we extended the primary study outcome (diagnosis of liver cirrhosis) to include liver-related death too (composite study outcome). Liver-related death was defined as one with a liver-related leading cause of death (**Appendix Table A4**). Given that the leading cause of death in DAR has been recorded since January 1, 2002, sensitivity analysis covered the period 2002-2018. Specifically, we confined the study population to incident IBD cases during 2002-2018 (along with their matched non-IBD individuals) and followed them up until diagnosis of cirrhosis/liver-related death (composite study outcome), death (leading cause other than liver-related), emigration, or end of follow-up (December 31, 2018), whichever came first. Persons in the matched non-IBD population were additionally censored in case of an IBD registration during follow-up, similarly to the primary analysis.

| Table A1. Diagnosis of cirrhosis | | | | |
| --- | --- | --- | --- | --- |
| Code | Description | | Type of Code | Source |
| *Compensated cirrhosis* | | | | |
| 571 | cirrhosis of liver | | ICD-8 | LPR |
| K70.3 | alcoholic liver cirrhosis | | ICD-10 | LPR |
| K74.2 | liver fibrosis with sclerosis | | ICD-10 | LPR |
| K74.4 | secondary biliary cirrhosis | | ICD-10 | LPR |
| K74.5 | biliary liver cirrhosis unspecified | | ICD-10 | LPR |
| K74.6 | other or unspecified liver cirrhosis | | ICD-10 | LPR |
| I85.9 | Esophageal varices without bleeding | | ICD-10 | LPR |
| I86.4 | Varicose veins in the stomach (Gastric varices, not bleeding) | | ICD-10 | LPR |
| I98.2 | Esophageal varices without bleeding in disease classified elsewhere | | ICD-10 | LPR |
| M49212 | cirrhosis | | SNOMED | pathology |
| M49500 | cirrhosis | | SNOMED | pathology |
| M49501 | incipient cirrhosis | | SNOMED | pathology |
| M49503 | cirrhosis with partially preserved architecture | | SNOMED | pathology |
| M49504 | active cirrhosis | | SNOMED | pathology |
| M49505 | cryptogenic cirrhosis | | SNOMED | pathology |
| M49506 | inactive cirrhosis | | SNOMED | pathology |
| M49510 | micronodular cirrhosis | | SNOMED | pathology |
| M49514 | active micronodular cirrhosis | | SNOMED | pathology |
| M49516 | inactive micronodular cirrhosis | | SNOMED | pathology |
| M49520 | macronodular cirrhosis | | SNOMED | pathology |
| M49524 | active macronodular cirrhosis | | SNOMED | pathology |
| M49526 | inactive macronodular cirrhosis | | SNOMED | pathology |
| M49527 | primarily macronodular cirrhosis | | SNOMED | pathology |
| M49528 | secondary macronodular cirrhosis | | SNOMED | pathology |
| M49530 | mixed micro- and macronodular cirrhosis | | SNOMED | pathology |
| M49570 | congestive cirrhosis | | SNOMED | pathology |
| M49580 | secondary biliary cirrhosis | | SNOMED | pathology |
| M49590 | primary biliary cirrhosis | | SNOMED | pathology |
| M49591 | primary biliary cirrhosis, noncirrhotic stage | | SNOMED | pathology |
| M49592 | primary biliary cirrhosis, cirrhotic stage | | SNOMED | pathology |
| M49660 | alcoholic cirrhosis | | SNOMED | pathology |
| M49690 | posthepatic cirrhosis | | SNOMED | pathology |
| *Decompensated cirrhosis* | | | | |
| 456.0 | varicose veins of other sites, of oesophagus | ICD-8 | | LPR |
| 785.3 | Ascites | ICD-8 | | LPR |
| I85.0 | Esophageal varices with bleeding | ICD-10 | | LPR |
| R18 | Ascites | ICD-10 | | LPR |
| K76.6 | portal hypertension | ICD-10 | | LPR |
| K76.7 | hepatorenal syndrome | ICD-10 | | LPR |
| M32601 | bleeding varicode veins | SNOMED | | pathology |
| F04440 | decompensation | SNOMED | | pathology |
| S74150 | portal hypertension syndrome | SNOMED | | pathology |

| Table A2. Etiology of liver disease | | | | | | |
| --- | --- | --- | --- | --- | --- | --- |
| Code | Description | | Type of code | | Source | |
| *Viral hepatitis* | | | | | | |
| 070 | infectious hepatitis | | ICD-8 | | LPR | |
| 999.2 | serum hepatitis | | ICD-8 | | LPR | |
| B15 | acute hepatitis A | | ICD-10 | | LPR | |
| B16 | acute hepatitis B | | ICD-10 | | LPR | |
| B17 | other acute viral hepatitis | | ICD-10 | | LPR | |
| B18 | chronic viral hepatitis | | ICD-10 | | LPR | |
| B19 | viral hepatitis without further specification | | ICD-10 | | LPR | |
| K73 | chronic hepatitis | | ICD-10 | | LPR | |
| *Immune-mediated* | | | | | | |
| K74.3 | (primary biliary cirrhosis)* | | ICD-10 | | LPR | |
| K75.4 | (autoimmune hepatitis) | | ICD-10 | | LPR | |
| K83.0F | primary sclerosis cholangitis | | ICD-10 | | LPR | |
| S63580 | autoimmune hepatitis | | SNOMED | | pathology | |
| S63605 | sclerosis cholangitis | | SNOMED | | pathology | |
| S63606 | primary sclerosis cholangitis | | SNOMED | | pathology | |
| S63607 | secondary sclerosis cholangitis | | SNOMED | | pathology | |
| *Primary metabolic* | | | | | | |
| 273.2 | haematochromatosis | | ICD-8 | | LPR | |
| 273.3 | Wilson’s disease (hepatolenticular degeneration) | | ICD-8 | | LPR | |
| E83.0B | Wilson’s disease | | ICD-10 | | LPR | |
| E83.1(A) | disturbances in iron metabolism (hemochromatosis) | | ICD-10 | | LPR | |
| E88.0A | alpha-1 antitrypsin deficiency | | ICD-10 | | LPR | |
| E88.0B | alpha-1 proteinase inhibitor deficiency | | ICD-10 | | LPR | |
| S10700 | alpha-1 antitrypsin deficiency disease | | SNOMED | | pathology | |
| S11920 | hemochromatosis | | SNOMED | | pathology | |
| S87250 | Wilson’s disease | | SNOMED | | pathology | |
| *Alcohol-related* | | | | | | |
| 291 | alcoholic psychosis | | ICD-8 | | LPR | |
| 303 | alcoholism | | ICD-8 | | LPR | |
| 577.10 | chronic alcoholic pancreatitis | | ICD-8 | | LPR | |
| E24.4 | alcohol-induced pseudo-Cushing’s syndrome | | ICD-10 | | LPR | |
| F10 | Mental disorders and behavioral disorders caused by the use of alcohol | | ICD-10 | | LPR | |
| F11 | Psychiatric disorders and behavioral disorders caused by the use of opioids | | ICD-10 | | LPR | |
| G31.2 | Degenerative changes in the nervous system caused by alcohol | | ICD-10 | | LPR | |
| G62.1 | Alcoholic polyneuropathy | | ICD-10 | | LPR | |
| G72.1 | Alcoholic myopathy | | ICD-10 | | LPR | |
| I42.6 | Alcoholic cardiomyopathy | | ICD-10 | | LPR | |
| K29.2 | Alcoholic gastritis | | ICD-10 | | LPR | |
| K70 | alcoholic liver disease | | ICD-10 | | LPR | |
| K85.2 | Acute alcoholic pancreatitis | | ICD-10 | | LPR | |
| K86.0 | Chronic alcoholic pancreatitis | | ICD-10 | | LPR | |
| T51.0 | Ethanol poisoning | | ICD-10 | | LPR | |
| T51.9 | Alcohol poisoning unspecified | | ICD-10 | | LPR | |
| X65 | Deliberate self-harm with alcohol | | ICD-10 | | LPR | |
| Z50.2 | Contact regarding rehabilitation after alcohol abuse | | ICD-10 | | LPR | |
| Z71.4 | Counseling and control for alcohol abuse | | ICD-10 | | LPR | |
| Z72.1 | Problem with alcohol consumption | | ICD-10 | | LPR | |
| S63590 | alcoholic hepatitis | | SNOMED | | pathology | |
| S63630 | toxic/drug-induced hepatitis | | SNOMED | | pathology | |
| S87720 | chronic alcoholism | | SNOMED | | pathology | |
| *Cryptogenic/NAFLD* | | | | | | |
| M45400 | | steatohepatitis | | SNOMED | | pathology |
| M50080 | | fatty degeneration | | SNOMED | | pathology |
| M50084 | | steatosis | | SNOMED | | pathology |
| M50085 | | macrivesicular steatosis | | SNOMED | | pathology |
| M50086 | | microvesicular degeneration | | SNOMED | | pathology |
| M50110 | | ballooning degeneration | | SNOMED | | pathology |
| M55200 | | fat deposition | | SNOMED | | pathology |
| M55280 | | fatty infiltration | | SNOMED | | pathology |

| Table A3. Diagnosis method of cirrhosis | | | |
| --- | --- | --- | --- |
| Code | Description | Type of code | Source |
| *Biopsy* | | | |
| P30610 | biopsy | SNOMED | pathology |
| P30611 | excisional biopsy | SNOMED | pathology |
| P30612 | electroexcision biopsy | SNOMED | pathology |
| P30613 | punch biopsy | SNOMED | pathology |
| P30614 | cryobiopsy | SNOMED | pathology |
| P30615 | endoscopic biopsy | SNOMED | pathology |
| P30616 | endoscopic brush biopsy | SNOMED | pathology |
| P30617 | abrasion/vabrasio | SNOMED | pathology |
| P30618 | endoscopic biopsy, imaging guided | SNOMED | pathology |
| P30619 | random biopsy | SNOMED | pathology |
| P3061A | incisional biopsy | SNOMED | pathology |
| P3061K | control biopsy | SNOMED | pathology |
| P3061X | donor biopsy | SNOMED | pathology |
| P30990 | needle biopsy | SNOMED | pathology |
| P30991 | needle biopsy, thin needle | SNOMED | pathology |
| P30992 | needle biopsy, coarse needle | SNOMED | pathology |
| P30993 | needle biopsy, medical indication | SNOMED | pathology |
| P30995 | needle biopsy and aspirate | SNOMED | pathology |
| P30996 | needle biopsy with imprint | SNOMED | pathology |
| P30997 | needle biopsy, CT-guided | SNOMED | pathology |
| P30998 | needle biopsy, coarse needle, CT-guided | SNOMED | pathology |
| P30999 | needle biopsy, stereotactic | SNOMED | pathology |
| P3099A | needle biopsy, MRI-guided | SNOMED | pathology |
| P3099B | needle biopsy, coarse needle, MRI-guided | SNOMED | pathology |
| P3099C | needle biopsy, endoscopic ultrasound guided | SNOMED | pathology |
| KJJA00 | exploration of liver | ICD-10 | LPR |
| KJJA10 | hepatotomy | ICD-10 | LPR |
| KJJA20 | liver biopsy | ICD-10 | LPR |
| KJJA21 | laparoscopic biopsy of liver | ICD-10 | LPR |
| KJJA23 | open needle biopsy of liver | ICD-10 | LPR |
| KJJA24 | laparoscopic needle biopsy of liver | ICD-10 | LPR |
| KJJA26 | transjugular liver biopsy | ICD-10 | LPR |
| *Imaging* | | | |
| *Ultrasound* | | | |
| UXUD | ultrasound examinations of the abdomen | ICD-10 | LPR |
| UXUD05 | ultrasound examinations of the abdomen, focused | ICD-10 | LPR |
| UXUD10 | ultrasound examinations of the upper abdomen | ICD-10 | LPR |
| UXUD11 | laparoscopic ultrasound examinations of the upper abdomen | ICD-10 | LPR |
| UXUD50 | endoscopic ultrasound examination of the liver | ICD-10 | LPR |
| UXUD52 | quantitative US/Doppler of the vessels of the liver | ICD-10 | LPR |
| UXUD70 | ultrasound examination of the liver | ICD-10 | LPR |
| *CT* | | | |
| UXCD | CT scans of the abdomen and pelvis | ICD-10 | LPR |
| UXCD00 | CT scan of the abdomen | ICD-10 | LPR |
| UXCD10 | CT scan of the upper abdomen | ICD-10 | LPR |
| UXCD15 | CT scan of the lower abdomen, incl. pelvis | ICD-10 | LPR |
| UXCD40 | CT scan of liver | ICD-10 | LPR |
| *MR* | | | |
| UXMD | MRI scans of the abdomen and pelvis | ICD-10 | LPR |
| UXMD10 | MR scan of the upper abdomen | ICD-10 | LPR |
| UXMD15 | MR scan of the lower abdomen, incl. pelvis | ICD-10 | LPR |
| UXMD40 | MRI scan of liver | ICD-10 | LPR |

| Table A4. Liver-related leading cause of death | | | |
| --- | --- | --- | --- |
| Code | Description | Type of code | Source |
| K700 | alcoholic fatty liver | ICD-10 | DAR |
| K701 | alcoholic hepatitis | ICD-10 | DAR |
| K702 | alcoholic fibrosis and sclerosis of liver | ICD-10 | DAR |
| K703 | alcoholic cirrhosis of liver | ICD-10 | DAR |
| K704 | alcoholic hepatic failure | ICD-10 | DAR |
| K709 | alcoholic liver disease, unspecified | ICD-10 | DAR |
| K710 | toxic liver disease with cholestasis | ICD-10 | DAR |
| K711 | toxic liver disease with hepatic necrosis | ICD-10 | DAR |
| K712 | toxic liver disease with acute hepatitis | ICD-10 | DAR |
| K717 | toxic liver disease with fibrosis and cirrhosis of liver | ICD-10 | DAR |
| K719 | toxic liver disease, unspecified | ICD-10 | DAR |
| K720 | acute and subacute hepatic failure | ICD-10 | DAR |
| K721 | chronic hepatic failure | ICD-10 | DAR |
| K729 | hepatic failure, unspecified | ICD-10 | DAR |
| K743 | primary biliary cirrhosis | ICD-10 | DAR |
| K746 | other and unspecified cirrhosis of liver | ICD-10 | DAR |
| K750 | abscess of liver | ICD-10 | DAR |
| K754 | autoimmune hepatitis | ICD-10 | DAR |
| K758 | other specified inflammatory liver diseases | ICD-10 | DAR |
| K761 | chronic passive congestion of liver | ICD-10 | DAR |
| K766 | portal hypertension | ICD-10 | DAR |
| K769 | liver disease, unspecified | ICD-10 | DAR |

| ***Table A5*. Liver disease (LD) registration(s) among persons with cirrhosis in the study population (N=2,741)** | | | | | | | | |
| --- | --- | --- | --- | --- | --- | --- | --- | --- |
| **LD indication I** | |  | **LD indication II** | |  | | **LD indication(s) III** | |
| **No. of persons (C %)** | |  |  |  |  | **No. of persons (C %)** | | |
| Viral hepatitis | 166 (6.1) |  | Viral hepatitis only | 74 (2.7) |  | Viral hepatitis only | | 74 (2.7) |
| Immune-mediated | 78 (2.8) |  | Viral hepatitis + | 92 (3.3) |  | Immune-mediated only | | 66 (2.4) |
| Primary metabolic | 14 (0.5) |  | Immune-mediated only | 66 (2.4) |  | Primary metabolic only | | 8 (0.3) |
| Alcohol-related | 1,070 (39.0) |  | Immune-mediated + | 12 (0.4) |  | Alcohol-related only | | 1,032 (37.6) |
| NAFLD* | 1,413 (51.6) |  | Primary metabolic only | 8 (0.3) |  | At least two types | | 148 (5.4) |
|  |  |  | Primary metabolic + | 6 (0.2) |  | NAFLD* | | 1,413 (51.6) |
|  |  |  | Alcohol-related only | 1,032 (37.6) |  |  | |  |
|  |  |  | Alcohol-related + NAFLD | 38 (1.4) |  |  | |  |
|  |  |  | NAFLD | 45 (1.6) |  |  | |  |
|  |  |  | No registration | 1,368 (50.0) |  |  | |  |

*NAFLD as per registration of specific code or absence of any registration at all.

C: column; +: at least one additional indication below the reported one (according to the suggested hierarchy) – e.g. ‘viral hepatitis +’ means viral hepatitis and at least one registration of immune-mediated, primary metabolic, alcohol-related and/or NAFLD.
